# Systematic Review of Hybrid Vision Transformer Architectures for Radiological Image Analysis

**DOI:** 10.1101/2024.06.21.24309265

**Authors:** Ji Woong Kim, Aisha Urooj Khan, Imon Banerjee

## Abstract

**Background:** Vision Transformer (ViT) and Convolutional Neural Networks (CNNs) each possess distinct strengths in medical imaging: ViT excels in capturing long-range dependencies through self-attention, while CNNs are adept at extracting local features via spatial convolution filters. However, ViT may struggle with detailed local spatial information, critical for tasks like anomaly detection in medical imaging, while shallow CNNs may not effectively abstract global context.

**Objective:** This study aims to explore and evaluate hybrid architectures that integrate ViT and CNN to leverage their complementary strengths for enhanced performance in medical vision tasks, such as segmentation, classification, and prediction.

**Methods:** Following PRISMA guidelines, a systematic review was conducted on 28 articles published between 2020 and 2023. These articles proposed hybrid ViT-CNN architectures specifically for medical imaging tasks in radiology. The review focused on analyzing architectural variations, merging strategies between ViT and CNN, innovative applications of ViT, and efficiency metrics including parameters, inference time (GFlops), and performance benchmarks.

**Results:** The review identified that integrating ViT and CNN can mitigate the limitations of each architecture, offering comprehensive solutions that combine global context understanding with precise local feature extraction. We benchmarked the articles based on architectural variations, merging strategies, innovative uses of ViT, and efficiency metrics (number of parameters, inference time(GFlops), performance).

**Conclusion:** By synthesizing current literature, this review defines fundamental concepts of hybrid vision transformers and highlights emerging trends in the field. It provides a clear direction for future research aimed at optimizing the integration of ViT and CNN for effective utilization in medical imaging, contributing to advancements in diagnostic accuracy and image analysis.

**Summary Statement:** We performed systematic review of hybrid vision transformer architecture using PRISMA guideline and perfromed through meta-analysis to benchmark the architectures.

**ACM Reference Format:** Ji Woong Kim, Aisha Urooj Khan, and Imon Banerjee. 2018. Systematic Review of Hybrid Vision Transformer Architectures for Radiological Image Analysis. *J. ACM* 37, 4, Article 111 (August 2018), 16 pages. https://doi.org/XXXXXXX.XXXXXXX

## 1 INTRODUCTION

Convolutions Neural Networks (CNN) were capable of learning inductive bias and were the state-of-the-art for many medical imaging applications [2]. Recently, vision Transformer has been adopted in a lot of applications in medical imaging domain and demonstrated comparative performance [12, 25, 34]. Since, the transformer has strength in capturing the long-range dependencies by using self-attention mechanism it has shown a great performance for complex natural language processing tasks[17, 42]. To identify small anomalies in anatomical imaging, the local correlation among the neighboring pixels also matters in addition to the long-range dependencies [40]. CNN architecture is known for being capable to capture such local dependencies[15, 36, 37]. Thus the Vision Transformer and CNN have complimentary strengths to process medical imaging for various applications, e.g. segmentation, classification, prediction. Therefore, many works have been attempting to combine these two architectures to have strength to capture both global and local contents of the images as hybrid concept architectures[22].

While CNN with the help of spatial convolution filters can primarily learn local features, shallow CNN networks with less layers often struggle to understand the global context in the image given the limitation of abstraction. In contrast, ViT learns the long-range dependencies via self-attention between the image patches to understand the global context while the patch based positional encoding mechanism may miss relevant local spatial information and it usually cannot attain the performance of CNNs on small-scale dataset. This limitation of ViT has been highlighted in multiple recent studies in particularly radiology domain where the findings are minute and contain within a small spatial location [24, 29]. Combining CNN and ViT in a hybrid modeling architecture can overcome the limitations of both and ultimately provide an opportunity to learn the global and local spatial context in an end-to-end model. Given many attempts to embrace strength of both ViT and CNN in radiology domain in a single end-to-end framework, we conducted the systematic review to understand various trends of how the Hybrid Vision Transformer (CNN+ViT) was used to address image processing challenges in radiology and create a comprehensive benchmarking to guide future development. Shamshad et. al. [34] recently published a comprehensive survey on transformer applications for medical images; however the survey was primarily focused on the broad transformer architectures and only briefly mentioned the hybrid structures’ pros and cons. while we primarily focus on benchmarking the hybrid architectures for radiological images.

Based on the PRISMA guideline, we have selected 28 published articles for full text review that proposed novel hybrid architecture for medical vision tasks. We performed three-level meta-analysis (a) overall architectural variations for the design of the hybrid CNN and ViT; (ii) merging strategies between CNN and ViT; (iii) purpose of innovative ViT usages; and (iv) design efficiency of the architectures in terms of number of parameters and inference time efficiency. To the best of our knowledge, there exist no systematic review paper that focuses on the hybrid architectures that combine ViT and CNN for the usage in radiology domain. While there exists survey papers that analyze ViT or CNN usage separately in medical imaging domain [34], we particularly focused on how the ViT is combined with other modules as a hybrid vision transformer architecture and applied to analyze varying scale features in radiology images. Through the meta-analysis, we not only analyzed and summarized the contents of the published literature, our also paper defines fundamental concepts of the hybrid vision transformer to make clear pathway for future research in this area.

## 2 METHOD

### 2.1 Study Selection

Figure 1 outlines the search strategy. Initially, papers for review were identified from Google Scholar, encompassing all articles published between Jan, 2020 and Aug, 2023. The first screening pass identified papers pertaining to six keywords [(‘Vision transformer’, ‘ViT’, ‘Hybrid ViT’) AND (‘Radiology’, ‘Image Analysis’, ‘Modeling’)]. Papers were excluded based on: (i) publication year before 2020 and after 2023 Aug, (ii) not peer-reviewed, and (iii) proposed usage not related to radiology.

**Fig. 1.**
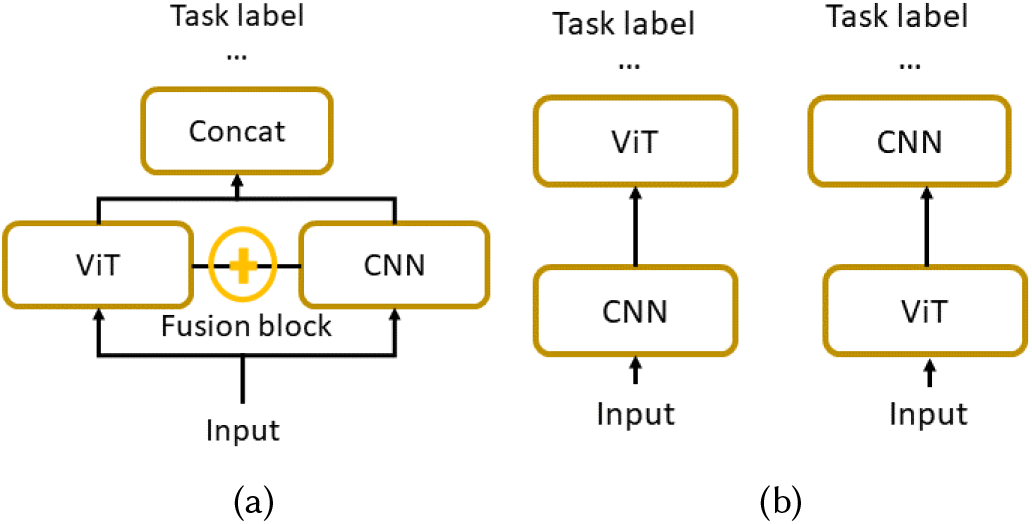
Architecture variations-(a) parallel and (b) sequential

**Fig. 2.**
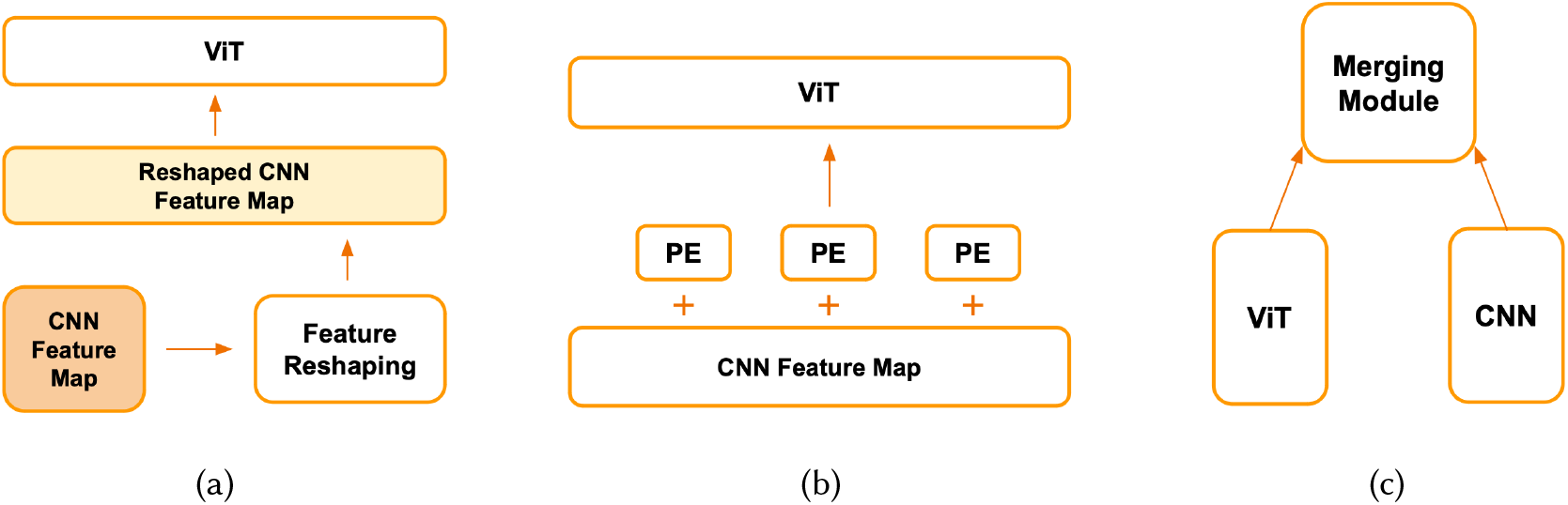
Merging Strategies variations - (a) Feature reshaping, (b) Positional encoding, and (c) Fusing Module.

To properly filter papers, we parsed the methods and experiments sections to determine the architecture and usage. We selected only architectures that combine self-attention or transformer modules in visual feature extraction and integrate with CNN modules. Vanilla ViT also proposed CNN embedding for input patches [28], thus we defined hybrid ViT as the Vision Transformer architecture that integrates CNN more extensively in end-to-end learning than the variant of vanilla ViT. Additionally, hybrid transformer architectures that used transformer modules solely for text feature extraction were excluded, as we focused on transformer usage for visual radiologic features.

Finally, papers that only concatenated the output of two architectures without proposing any new design modifications were excluded. Subsequently, three independent reviewers, JK, AU, and IB, scanned the retrieved papers and selected relevant papers based on predefined inclusion and exclusion criteria. Conflicts were resolved by majority voting.

### 2.2 Meta-Analysis

We primarily focused on four meta analysis factors as described below.

#### 2.1.1 Architecture

We categorized the existing hybrid architectures into parallel and sequential design (Fig. 1). Parallel design includes the one where CNN and ViT modules used in-parallel to cooperatively learn the feature representation from the data and various intermediate fusion functions is generally used to increase the co-understanding between the modules, such as cross-attention. Note that both modules are parsing the data in the same level of granularity and have less dependence on each other. In contrast, CNN and ViT are used in sequence in sequential design where output of one module is directly passed onto others. One module is used for initial feature extraction and other module for generating an abstract view based on previous module interpretation. Therefore, the dependency between the CNN and ViT modules is much higher in the sequential design.

#### 2.2.2 Merging Strategy

Based on the architectural variations (sequential or parallel), various strategies have been employed to merge the CNN and ViT to effectively utilize their respective strengths. We have identified three broad techniques for merging ViT and CNN: (i) *feature reshaping* this is extremely well adopted technique for the sequential architecture where output of one module is resized using simple linear function (such as flatting) to be fed into the other. For example, given that ViT only parse sequential input tokens, CNN feature maps are flattened into sequential tokens if the output is passed to ViT; (ii) *positional encoding* - Since flattened feature maps lose spatial information, often architecture that use feature reshaping also include positional encoding to capture the spatial information based on the feature map; (iii) *fusing module* - for the parallel design where both of the modules are co-learning together, most of the literature used linear combination of ViT and CNN output to create a fusion between these two modules or use a different model for learning the fusion parameters.

#### 2.2.3 Transformer utilization

Given the usage of transformer varied widely between the hybrid models, we also categorized the hybrid model based on the final usage of the transformer - (i) *Encoder* - when the transformer is used to embed the raw or processed data. In other words generate a compressed representation of input; (ii) *Decoder* - when the transformer is used for the generation of reconstruction or interpretation.

#### 2.2.4 Application

We observed that hybrid transformer architectures are adopted in various problems of computer vision for healthcare, starting from classic tasks like classification, segmentation, reconstruction, registration to regression, synthesis, view combination, and text generation. We aimed to group the designs based on the architecture to better highlight the design choices and metric for performance evaluation.

### 2.3 Benchmarking criteria

We used five criteria to benchmark these models which are centered around understanding the utility and efficiency of the hybrid models.

#### 2.3.1. Modality

Hybrid models are primary developed to deal with high dimensional spatial data (e.g. 2D, 3D, 4D) to process them in computationally efficient way through transformer while keeping both local and global spatial context. Therefore, we consider the imaging modality (2D - X-ray, 2D + time - Ultrasound (US), 3D - Magnetic Resonance (MR), Computed Tomography (CT), Positron Emission Tomography (PET)) as a primary benchmarking criteria which is ideally a proxy to represent the data dimensionality.

#### 2.3.1. Model size

We measured the model size by the number of trainable parameters. In theory, as the number of trainable parameters increases, so does the need for more training samples [11], which limits the large model’s applicability for interesting clinical use cases due to the lack of training data and manual annotation in the clinical domain. However, the number of training parameters also depends on the input data dimension. Therefore, we defined an efficient hybrid design as a model that can handle high dimensional input data with fewer training parameters. We primarily record the number of trainable parameters either from the published paper (if documented) or by loading the model summary in python, if the model is supplied by the authors with academic open-source license.

#### 2.3.3. Computing efficiency

To measure the computing efficiency, we used the flops/image as a benchmarking creteria to demonstrate feasibility of applying the algorithm real time. Flops denotes the number of floating point operations performed to run the inference on a single image. Given the assumption that the hardware configurations may vary, flops shows a standardize hardware independent measure of the algorithmic efficiency of the hybrid model. Though, we calculated and tested all the models on 4 A100 GPU cores with 32GB memory. Similar to model size, we rely on the published manuscript for the flops if documented or calculated the flops by doing inference on a single random input image generated based on the dimension specified by the authors.

#### 2.3.4. Training data size

Though training sample size is not directly related with the hybrid model design, we incorporated it as a benchmarking creteria to highlight task and modality specific trend of the targeted use cases and how it ultimately affects the model performance. We directly capture the number of training samples from the reported documentation by the original authors. If multiple training setting was mentioned in the paper, we only selected the largest cohort setting and report the number of total exams included.

#### 2.3.5. Performance

Based on the applications (Sec. 2.2.4), we ultimately benchmark the reported performance. Given the issue of not availability of the shared codebase or a common dataset, we only documented the reported performance by the original authors on distinct datasets. However, the performance benchmark highlights overall performance for a task based on a standard task-specific metric, e.g. Dice for segmentation accuracy, AUC for classification prediction true positive and false positive trade-off at different probability threshold (F1 if AUC is not reported), Structural Similarity Index (SSIM) for reconstruction and registration quality compared to the input data, and Bleu1 score for evaluating the quality of text generation compared to the original reference text.

## 3 RESULTS

In Fig. 3, we present the PRISMA diagram with total articles included and excluded at each step of filtering. Finally, within the scope of this survey, we analyzed 28 articles that satisfied all our inclusion criteria. Table 1 shows the meta analysis results and Table 2 benchmarking according to the defined meta analysis and benchmarking criteria in Sec. 2.2 and Sec. 2.3 respectively.

**Table 1.** Meta-Analysis of the existing hybrid architectures performed based on the criteria defined in Sec.2.2. Studies are grouped based on the targeted applications. ‘Transf.’, ‘Util.’ refers to Transformer and Utility respectively.

| Reference | Application | Architecture | Merging | Transf. Util. | Transf. Backbone | CNN Backbone |
| --- | --- | --- | --- | --- | --- | --- |
| U-net Transformer[31] | Segmentation | Sequential | Fusing | Encoder | Multi-head Cross-Attention | U-Net |
| UTNet[15] |  | Sequential | Feature Reshaping<br>Positional Encoding | Encoder<br>Decoder | Multi-head Self-Attention | U-Net |
| CPT U-Net[48] |  | Parallel | Fusing | Encoder<br>Decoder | Pyramid Vision Transformer | U-Net |
| UNETR[19] |  | Sequential | Feature Reshaping | Encoder | Vision Transformer | U-Net |
| Swin UNETR[18] |  | Sequential | Feature Reshaping<br>Fusing | Encoder | Swin Transformer | U-Net |
| COTRNet[35] |  | Sequential | Feature Reshaping<br>Positional Encoding | Encoder | Light Vision Transformer | U-Net |
| Cotr[43] |  | Sequential | Feature Reshaping<br>Positional Encoding | Encoder | Deformable Transformer-encoder | U-Net |
| Hybrid ViT and CNN[39] |  | Sequential | Fusing | Encoder<br>Decoder | Vision Transformer | U-Net |
| TransBTS[40] |  | Sequential | Feature Reshaping<br>Positional Encoding | Encoder | Light Vision Transformer | U-Net |
| Bitr U-Net[21] |  | Sequential | Feature Reshaping<br>Positional Encoding | Encoder | Vision Transformer | U-Net<br>CBAM |
| After U-Net[44] |  | Sequential | Feature Reshaping | Encoder | Axial Fusion Transformer | U-Net |
| WAU [26] |  | Sequential | Feature Reshaping | Decoder | Window Attention | Group Convolution<br>Depthwise Separable CNN |
| ChexVit[13] | Classification | Sequential | Feature Reshaping<br>Positional Encoding | Encoder | Vision Transformer | CheXNet [32] |
| TECNN[1] |  | Parallel | Feature Reshaping<br>Fusing | Encoder | Vision Transformer | DenseNet |
| SLATER[23] | Reconstruction | Sequential | Feature Reshaping<br>Positional Encoding | Decoder | Cross-Attention | Specialized CNN |
| 3D Transformer GAN [27] |  | Sequential | Feature Reshaping<br>Positional Encoding | Encoder<br>Decoder | Vision Transformer | Specialized CNN |
| T2Net [14] | Reconstruction | Sequenital | Feature Reshaping | Encoder | Task-Attention<br>Soft-Attention | Specialized CN |
| MIST-Net[30] |  | Sequential | Feature Reshaping<br>Fusing | Decoder | Swin Transformer<br>Reconstruction Swin-transformer | Specialized CNN |
| Ultrasound ViT[33] | Regression | Sequential | Feature Mapping | Encoder | Bert | ResNetAE/DenseNet |
| DTN[46] | Registration | Sequential | Feature Reshaping<br>Fusing | Encoder<br>Decoder | Dual Transformer | Specialized CNN |
| ViT-V-Net[5] |  | Sequential | Feature Reshaping<br>Positional Encoding | Encoder | Vision Transformer | Specialized CNN |
| Transmorph [4] |  | Sequential | Feature Reshaping | Encoder | Swin Transformer | U-Net |
| ResViT[10] | Image Synthesis | Sequential | Feature Reshaping | Encoder | Vision Transformer | Specialized CNN |
| TransCT[47] | Restoration | Parallel | Feature Reshaping | Encoder<br>Decoder | Vision Transformer | Specialized CNN |
| Multi-View ViT[38] | Combining Views | Sequential | Feature Reshaping | Encoder | Cross View-Attention | ResNet |
| AlignTransformer[45] | Report Generation | Sequential | Feature Reshaping | Encoder | Align Hierarchical-Attention | ResNet |
| R2Gen[7] |  | Sequential | Feature Reshaping | Encoder<br>Decoder | Vision Transformer | Pretrained (ResNet, VGG) |

**Table 2.** Benchmark Table. See Sec. 2.3 for benchmarking creteria definition. Inaccessibility of the models are marked as ‘–’ which restrict to calculate the benchmarking creteria.

| Reference | Modality | Parameters(M) | Inference Time(GFLOPs) | Sample Size | Performance |
| --- | --- | --- | --- | --- | --- |
| U-net Transformer[31] | CT | 42.5 | – | TCIA (public):82 total | Dice:78.5 |
| UTNet[15] | MR | 14.4 | 40.9 | M&Ms [3](public)<br>Training:150/Test:200 | Dice:88.3 |
| CPT U-Net[48] | CT | 123.8 | 150.6 | Synapse<sup>1</sup>(public) 30 | Dice 0.81 |
| UNETR[19] | CT/MRI | 92.5 | 41.1 | BTCV:20subjects,<br>MSD:484 CT/MRI | Dice:0.89 |
| Swin UNETR[18] | MRI | 61.9 | 394.8 | BraTS21<br>Training:1251 / Val:219 | Dice:0.92 |
| COTRNet[35] | CT | – | – | Kits21<br>Training:240 / Val:60 | Dice:0.61 |
| Cotr[43] | CT | 41.9 | 399.2 | Kits21<br>Training:240 / Val:60 / Test:100 | Dice:0.61 |
| Hybrid ViT and CNN[39] | CT | – | – | Synapse<br>Training:18 / Val:12 | Dice:0.87 |
| TransBTS[40] | MRI | 32.9 | 333.0 | BraTS19<br>Train:335 / Val:125 | Dice:0.9 |
| TransUNet[6] | CT/MRI | 105.1 | 1186.9 | Synapse<br>Train:18 / Val:12 | Dice: 0.77 |
| Bitr U-Net[21] | MRI | 43.4 | 186.2 | BraTS21<br>Training:1251 / Val:219 | Dice:0.92 |
| After U-Net[44] | CT | 41.5 | – | BCV Training:18 / Test:12 | Dice:0.81 |
| WAU[26] | CT/MRI | 21.8 | 15.94 | Synapse: 18<br>MSD: 484 | Dice:0.80 |
| ChexVit[13] | X-Ray | – | – | ChestX-Ray14 [41]<br>Training:86,524 / Val:25,596 | AUC:0.83 |

<sup>1</sup>https://www.synapse.org/#Synapse:syn3193805/wiki/89480

|  |  |  |  |  |  |
| --- | --- | --- | --- | --- | --- |
| TECNN[1] | MRI | 22.5 | – | BraTS, Figshare<br>Training:998 + / Val:285 + / Test: 142 + | Recall:0.97<br>Precision:0.967<br>F1-Score:0.968 |
| SLATER[23] | MRI | 36.0 | 174.8 | IXI:Training:25 / Val:5 / Test:10 | SSIM(T2): 97.77 |
| 3D Transformer GAN [27] | PET | 42.0 | – | Brain MRI<br>Training:10935(15subjects×729patches)<br>Val:Leave-One-Out-Cross-Validation | SSIM:0.986 |
| T2Net [14] | MRI | 1.4 | 140.3 | IXI Dataset, Clinical MRI Dataset<br>Training:420 / Val:60 / Test:120 | SSIM:0.87 |
| MIST-Net[30] | CT | 11.8 | 576.0 | 2016 NIH AAPM Mayo Challenge<br>Training:4,274 sinograms<br>Test:391sinograms | SSIM:0.98 |
| Ultrasound ViT[33] | Echocardiogram | 346.8 | 521.7 | Echonet-Dynamic<br>Training:7522 / Val:1504 / Test:1504 | MAE:6.77 |
| DTN[46] | MRI | – | – | Oasis, IXI, BraTS<br>Training:256 / Val:19 / Test:150 | Dice:0.76 |
| ViT-V-Net[5] | MRI | 31.5 | 778.4 | In-House T1 Weight<br>Training:182 / Val:26 / Test:52 | Dice:0.72 |
| Transmorph[4] | CT/MRI/XCAT | 46.7 | 1427.0 | Oasis<br>Training:256 / Val:19 / Test:150 | Dice:0.816 |
| ResViT[10] | CT/MRI | 123.4 | 973.0 | IXI, BraTS, CT-MRI<br>Training:59 / Val:29 / Test:42 | SSIM<br>T1,T2->FLAIR:0.886 |
| TransCT[47] | CT | 12.6 | 598.7 | 2016 NIH AAPM Mayo Challenge<br>Training:7patients / Val:1 / Test:2 | SSIM<br>0.92 |
| Multi-View ViT[38] | X-Ray | 23.6 | 9.5 | CheXpert<br>Training:23628(16810 patients)<br>Val:3915/ Test:3870 | AUC<br>0.834 |
| AlignTransformer[45] | X-Ray | – | – | MIMIC CXR<br>Training:368,960<br>Val:2,991<br>Test:5,159 | BLEU1<br>0.378 |
| R2Gen[7] | X-Ray | 78.4 | 35.4 | MIMIC CXR<br>Training:368,960<br>Val:2,991<br>Test:5,159 | BLEU1<br>0.353 |

**Fig. 3.**
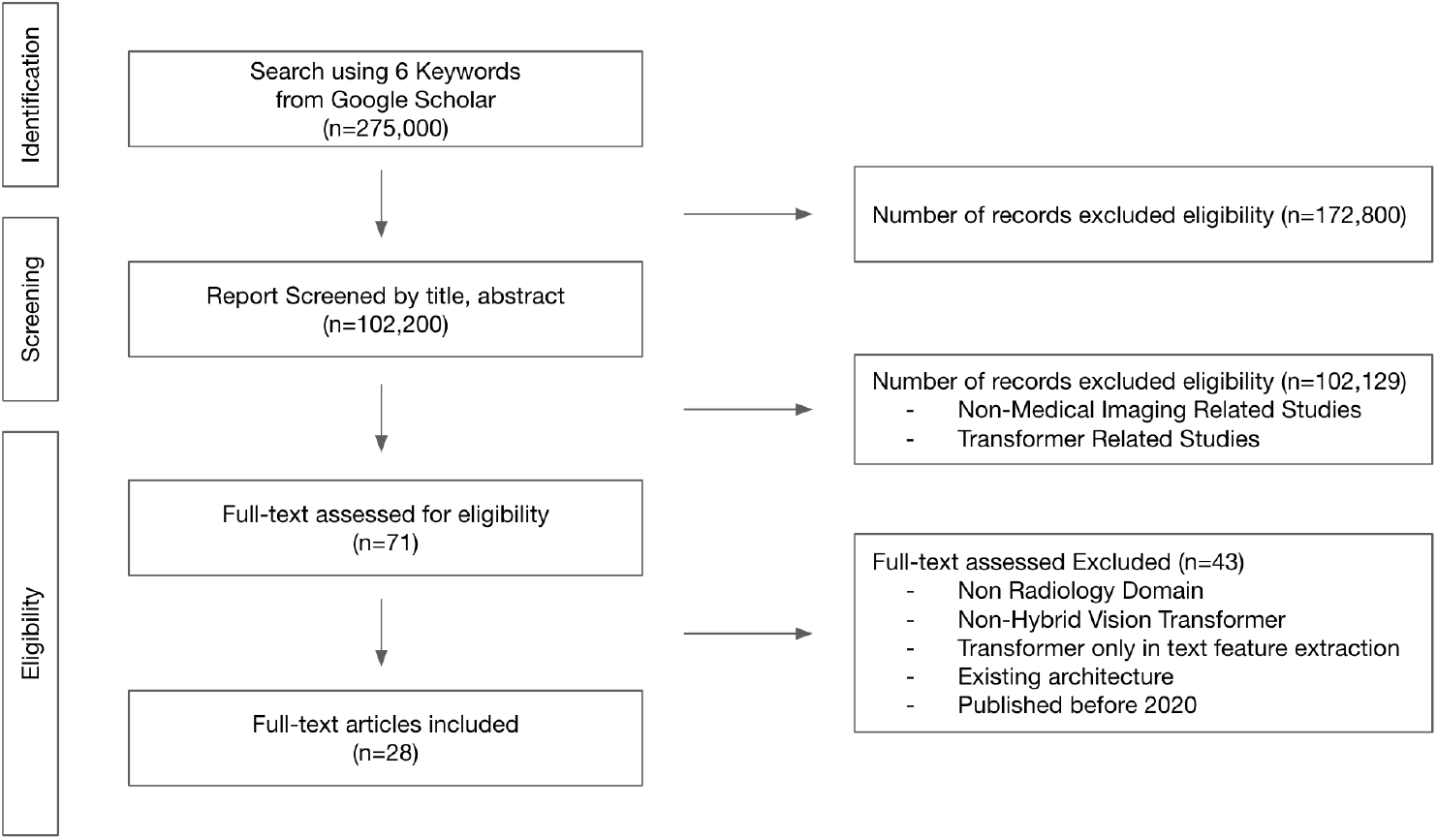
PRISMA study selection diagram for Hybrid Vision Transformer in Radiology

### Meta-analysis: Architecture

Despite parallel architecture enhances cooperative learning between CNN and ViT, as simplistic design option, most hybrid architectures follow sequential structure (25 out of 28) where the output of the CNN feature extractor block is fed into ViT for generating compressed representation (Table 1). Particularly, the hybrid models proposed for segmentation mostly (11 out of 12) follow the U shape architecture based on U-Net, and transformer with self-attention module is included between encoder and decoder branch within the U-Net shape backbone. For the classification, restoration and reconstruction, similar sequential architecture idea has been adopted where CNNs are used as feature extractor and feature maps are flattened and fed into transformer with positional encoding which either calculated in the feature space [13] or referred to the image space using learnable parameters [40]. After the transformer encoding, CNN decoder is included based on the image generation task requirement, e.g. registration, reconstruction.

Parallel architectures were particularly proposed to conserve same level features using both CNN and transformer; however fusing the feature spaces at multiple level is a challenging problem and needs innovative measure. For segmentation, CPT U-Net [48] adopted the CNN with parallel branch with ViT and fuse the each feature maps from parallel branches. CPT U-Net shows that performance also varies depending on the modification of fusing module. For the parallel classification paradigm, TECNN [1] utilizes parallel architecture using transfomer pathway where the CNN feature maps are flattened at each step and fed into the transformer, and ultimately merge module combines both feature space and compute classification score using softmax. As parallel architecture for high quality image restoration, TransCT decomposed the images into high (HF) and low (LF) frequency component and used CNN to parse LF and transformer to parse HF. To combine the transformer and CNN features, they again utilize a simplistic ResNET (2 Conv Layers) to get the final output.

### Meta-analysis: Merging Strategy

As also highlighted by the dominance of sequential architectures (25 out of 28), irrespective of the downstream application, most of existing hybrid architectures [13, 15, 27, 35, 40] leverage the long-context learning ability of the transformers after calculating feature maps from CNN by reshaping the feature maps and by adding the positional encoding. Separate fusing modules are primarily leveraged by the parallel architectures where either linear merging or another model is used for fusing the features from transformer and CNN [1, 39]. Interstingly, being a sequential network, DTN [46] simultaneously leverage the feature reshaping for handling exchange between CNN and transformer, and fusion module for aggregation of two parallel transformer blocks for temporal image frames. TransMorph [4] designed an unique strategy which we grouped under the broad category of feature reshaping, though they developed a patch merging module to reshape and align the features between the 3D Swin transformer blocks in the encoder and the feature maps generated at each resolution are sent into a ConvNet decoder to produce an output.

### Meta-analysis: Transformer utilization

Transformers are solely used for additional compression of the bottleneck features to capture the global context - out 18 of 28 studies. Interestingly, 7 studies applied transformer for both encoding and decoding purposes - particularly for target image generation tasks, such as segmentation, registration, restoration. Image to generation models (R2GEN [7]) primarily use transformer for the language generation task where CNN is being used as a image feature extractor. A compelling use of transformer is observed in 3D transformer GAN which devices a 12 layers transformer network between the encoderCNN and decoderCNN where 6 transformer layers performs encoding and 6 layers performs decoding before feeding into the CNN decoder block. The primary different from the original transformer, the transformer block applies parallel decoding to achieve parallel sequence prediction that is reshaped and processed by 1×1×1 convolution.

### Meta-analysis: Application

The hybrid architectures are adopted for all fundamental medical image analysis tasks, e.g. segmentation (12), classification (2), reconstruction (4), registration (3). Additionally, we observed some innovative applications of the hybrid architectures. For example, ResViT [10] leveraged the contextual sensitivity of vision transformers along with the precision of convolution operators to generate missing multi-Contrast MR series and compared against state-of-art convolution only GAN based models and showed that hybrid methods outperformed. TransCT [47] improvise a to enhance the final CT image quality from a low dose CT images by using content features from transformer and latent texture features from CNN. Multi-View ViT [38] designed a cross-view transformer to transfer information between unregistered image views at the level of spatial feature maps, and validated for mammogram and chest X-ray.

### Benchmarking

Based on the five benchmarking criteria described in Sec. 2.3, we compared the existing 28 architectures. We observed that most the hybrid architectures (23/28) are applied for the high dimensional imaging modalities, such as MR, CT, PET. This deign choice could be influenced by the capability of the transformer-CNN hybrid to digest both local and global context from a high dimensional image space and generate a denser representation compared to CNN only encoder. Highest number of trainable parameters (346.8M) is observed in Ultrasound ViT[33] that process variable length echo cardiogram videos since they utilized 16 parallel BERT encoders for spatio-temporal reasoning and two parallel regression tasks where they used ResNetAE to distil the US frames into smaller dimension embedding (1024D) and the resulting embeddings are stacked for the clip, and BERT encoders are used to process variable length videos. T2Net[14] contains lowest number of trainable parameters (1.4M) where they proposed a multitask learning framework where shared parallel network backbone, leveraging knowledge from one task to speedup the learning of the other and increase flexibility for sharing complimentary features. Only CNN based model, such as DenseNet121 has 8.1M and ResNet-50 is approximately 25.6M, which shows the fact the hybrid architecture often does not increase the trainable parameter set as it helps to generate dense representation.

Theoretically, light-weight models with less parameters are faster during inference with less number of floating point operations; While T2Net has the lowest parameters, Multi-view ViT [38] for chest X-ray has the fastest inference speed which could be based on the fact that it process 2D compress X-ray images. Inference speed for segmentation (TransUNet [6]) and reconstruction (Transmorph [4]) models is often higher than the classification due to their pixel or patch-wise processing strategies. Following the similar trend as CNN, hybrid models are often trained and validated on open-source datasets, e.g. BraTS [16], Figshare [8], KiTS21 [20], TCIA [9], Synapse, with limited samples of high dimensional medical images, and obtained current state-of-the-art performance for all major medical image analysis tasks by outperforming its CNN or transformer only counterparts. This trends for all major medical image analysis tasks shows the benefit of designing hybrid architectures by combining both CNN and transformers.

## 4 DISCUSSION

Following PRISMA guideline, we performed a systematic review for the 28 hybrid CNN and transformer architectures for radiological image analysis task. The diverse roles of transformers, from encoding raw data to decoding generated outputs, highlight their versatility in medical image analysis tasks. While transformers excel at capturing long-range dependencies, their integration into hybrid architectures introduces challenges in balancing computational efficiency and performance across different applications. The wide range of applications for hybrid vision transformer architectures underscores their potential to address various challenges in radiology, from segmentation and classification to reconstruction and registration. Innovative approaches, such as missing image generation and image quality enhancement, demonstrate the versatility and adaptability of these models to clinical needs.

The predominance of sequential architectures in hybrid vision transformer models suggests a preference for a structured flow of information from CNN feature extraction to transformer-based representation learning. This sequential approach aligns well with the nature of medical image analysis tasks, where hierarchical feature extraction and global context understanding are crucial. The utilization of feature reshaping and positional encoding reflects efforts to bridge the gap between CNN and transformer representations, enabling effective fusion of spatial and contextual information. While sequential architectures predominantly rely on these strategies, parallel architectures explore alternative fusion methods, emphasizing the need for further research into optimal merging techniques.

However, challenges remain in ensuring robustness, interpretability, and scalability for real-world applications. Further validation on diverse datasets and clinical scenarios is essential to establish their efficacy and reliability. While hybrid architectures offer advanced capabilities in feature representation and context modeling, their computational demands pose practical challenges, particularly in resource-constrained clinical environments. Optimizing model size, training efficiency, and inference speed is crucial for facilitating widespread adoption and deployment in clinical settings. Moreover, the reliance of hybrid architectures on large-scale annotated datasets raises concerns about data availability and quality, especially for rare diseases and specialized imaging modalities. Addressing data scarcity through data augmentation, transfer learning, and domain adaptation techniques is critical for generalizing model performance across diverse clinical scenarios.

Future research should explore novel hybrid architectures that leverage the complementary strengths of CNNs and transformers while addressing their inherent limitations. Investigating alternative fusion strategies, attention mechanisms, and network architectures could lead to more efficient and effective hybrid models tailored to specific clinical tasks and modalities. Comprehensive validation studies on diverse clinical datasets are essential to assess the generalization and robustness of hybrid vision transformer models across different imaging modalities and pathologies. Collaborations between clinicians, radiologists, and machine learning researchers are crucial for co-designing and evaluating models that meet clinical needs and standards. Use of distinct private datasets for evaluation and reporting their performance using different metrics makes the comparison between the models extremely challenging. Standardized benchmarking criteria and open-source dataset is needed for the model understanding. We believe that our review will set a proper benchmakring framework for the hybrid models.

Efforts to optimize model inference speed, memory efficiency, and hardware compatibility are essential for enabling real-time deployment of hybrid architectures in clinical workflows. Leveraging hardware accelerators, model compression techniques, and cloud-based inference services can facilitate seamless integration into existing radiology infrastructure.

As hybrid vision transformer models become increasingly integrated into clinical practice, attention must be paid to ethical and regulatory considerations, including patient privacy, data security, and algorithmic transparency. By addressing challenges related to model design, computational efficiency, data availability, and ethical considerations, these models can significantly impact patient care and healthcare delivery, paving the way for a future where AI-powered radiology transforms diagnosis, treatment, and patient outcomes.

*Limitation* Due to non-standardize performance reporting style in literature, we were unable to create a standard performance benchmark and models were also validated on distinct datasets. If the model is not available with open-source license, we were unable to calculate the inference speed and trainable parameters. We did make an effort to reach the corresponding authors but didn’t get the model access.

## 5 ESSENTIALS

1. Hybrid vision transformer architectures preserves both local and global spatial dependencies in radiological images.
2. Such architecture holds tremendous promise for advancing medical image analysis in radiology, offering superior performance, interpretability, and clinical relevance.
3. Comprehensive validation studies on diverse clinical datasets are essential to assess the generalization and robustness of hybrid vision transformer models.
4. Efforts to optimize model inference speed, memory efficiency, and hardware compatibility are essential for enabling real-time deployment.

## Data Availability

All data produced in the present work are contained in the manuscript

## Footnotes

1 https://www.synapse.org/#Synapse:syn3193805/wiki/89480

